# Age-based variability in the association between restraint use and injury type and severity

**DOI:** 10.1101/2022.04.06.22273536

**Authors:** Marco H. Benedetti, Kayleigh D. Humphries, Rachel Codden, Shraddha Sagar, Joseph A. Kufera, Lawrence J. Cook, Jesse Norris, Nikiforos Stamatiadis, Roumen Vesselinov, Motao Zhu

## Abstract

**Purpose:** Previous studies have shown elderly individuals receive less relatively less protection from seat belts against fatal injuries, however it is less clear how seat belt protection against severe and torso injury changes with age. We estimated age-based variability in seat belt protection against fatal injuries, injuries with maximum abbreviated injury scale greater than 2 (MAIS3+), and torso injuries.

**Methods:** We leveraged the Crash Outcome Data Evaluation System (CODES) to analyze binary indicators of fatal, MAIS3+, and torso injuries. Using a matched cohort design and conditional Poisson regression, we estimated age-based relative risks (RR) of the outcomes associated with seat belt use.

**Results:** Seat belts were highly protective against fatal injuries for all ages. For ages 16-30, seat belt use was associated with 66% lower risk of MAIS3+ injury (RR 0.34, 95% CI 0.30, 0.38), whereas for ages 75 and older, seat belt use was associated with 38% lower risk of MAIS3+ injury (RR 0.62; 95% CI 0.45, 0.86). The association between restraint use and torso injury also appeared to attenuate with age.

**Conclusions:** Seat belt protection against MAIS3+ and torso injury attenuated with age. We encourage that injury prevention continues to be tailored to vulnerable populations like the elderly.

## INTRODUCTION

Studies of bodily injuries in motor vehicle crashes (MVCs) show that older individuals are at a greater risk for severe and fatal injuries, and that a greater proportion of elderly individuals in MVCs sustain thoracic, chest, and other torso injuries [1-13]. Furthermore, seat belts may provide less protection against fatal injuries to elderly vehicle occupants due to greater frailty [14, 15]. and that elderly occupants may be more prone to torso injuries caused by seat belts. For example, Ekambaram et al. (2019) attributed higher rates of skeletal chest injuries in middle aged and elderly occupants to forces being transmitted to the chest through the seat belt [5].

The literature on seat belt effectiveness in preventing motor vehicle fatalities and injuries is extensive. Comprehensive reviews and meta-analyses of fatal injuries and major injuries show, unsurprisingly, that seat belts are highly protective [16, 17]. Previous studies have also estimated what protection, if any, seat belts provide against torso injuries, including thoracic, abdominal, spinal, and pelvic injuries [18-22]. Because seat belts are the most effective intervention for preventing MVC injury, and this protective effect against fatal injury has been shown to decrease with age, it is pertinent to estimate age-based variability in seat belt protection in other types of injuries, such as severe and torso injuries. Such analyses are scarce in the current literature. An analysis of crash reports from South Korea and found that those aged 65 and older who failed to wear a seat belt had higher odds of severe injury than their properly restrained counterparts, and that the effect size corresponding to seat belt protection was greater among those aged 65 and older compared to younger age groups [23]. However, to our knowledge no analogous study of severe injuries in the United States is available, and we are not aware of any study that estimates age-based variability in seat belt protection against torso injuries.

The Crash Outcome Data Evaluation System (CODES) is well-suited to address research questions about relationships between increasing age and seat belt efficacy. By linking crash data to hospital data, CODES provides detailed crash and injury information on a large sample of MVC occupants, overcoming the limitations other MVC data systems. CODES has great utility in a variety of traffic safety applications beyond the analysis of fatal MVCs, such as over-reporting of seat belt use [24], motorcycle helmet laws and brain injuries [25], and extremities injuries to bicyclists [26]. With a primary goal of better understanding how seat belt effectiveness against different injury types varies with age, the present study leverages CODES data to assess age-based variability in the associations between seat belt use and three types of injuries – fatal, severe, and torso – in motor vehicle crashes.

## MATERIALS AND METHODS

### Data Source

CODES was initiated in 1992 by the National Highway Traffic Safety Administration [27]. At the time, the primary data sources for studies of MVC outcomes in the United States were (i) FARS, a census of fatal MVCs; and (ii) police accident reports (PARs), both of which posed major limitations to inferring about non-fatal injuries. FARS data are limited to only fatal crashes by design. Although FARS is the gold standard data for fatal crashes, they exclude any crashes in which no occupant died, limiting their utility in studies of non-fatal injuries. On the other hand, while PAR data may provide a more complete set of injury crashes than FARS, police reported injuries may not accurately characterize true injury type and severity [28, 29].

CODES aimed to overcome the limitations of FARS and PAR data by probabilistically linking crash data, obtained via PARs, and hospital data from emergency departments, as well as inpatient and outpatient clinics [27, 30]. Probabilistic linkage is a method which uses properties of variables common to databases to determine the probability that two records refer to the same person and event and should be linked [31, 32]. CODES linkages are based on date of birth, sex, occupant type, date, time, and location using probabilistic record linkage via LinkSolv software [33]. To account for the uncertainty added by probabilistic linkage, the procedure is repeated five times, resulting in five data [30].

### Study Sample and Design

We leveraged CODES data from Ohio, Utah, Maryland, and Kentucky. We included all occupants aged 16 and older of passenger vehicles (e.g., sedans, SUVs, pickup trucks, minivans) with model year 2000 or newer that were involved in MVCs. We restricted the study period to span years 2009-2016, however Kentucky data were only available up to year 2014. Data were standardized according to the General Use Model format [30], allowing each site to share common variables.

To curtail potential confounding due to crash- and vehicle-level factors, we utilized a matched cohort design [34]. This study design is often used in studies of MVC injury [35-37] because it matches occupants in the same vehicles, thereby controlling for all crash- and vehicle-level factors. The use of a matched cohort design resulted in us creating three data sets, one for each of the three outcomes in our study (see the **Study Variables** subsection). These data sets were restricted to include only multi-occupant vehicles in which one or more of the occupants experienced the respective outcomes. Additional details on how the match cohort data were used to estimate relative risks are provided later in the **Statistical Analysis** subsection.

### Study Variables

We considered three binary outcome variables that were defined as follows:

1. Fatal Injury was equal to 1 when the hospital record or PAR indicates that the occupant died, and zero otherwise.
2. MAIS3+ Injury was equal to 1 when the occupant’s maximum abbreviated injury scale (MAIS) is equal to three or above, and zero otherwise [38]. Most injury scores were obtained from hospital diagnosis codes, however we also included occupants for whom no hospital injury data are available, but whose police-reported injury severity was fatal. Although we could have included those who were coded by police as having incapacitating injury, there is evidence that police-reported injuries do not align with diagnosed injuries, and that they tend to over-report severity [28, 29].
3. Torso Injury was equal to 1 when the occupant’s International Classification of Diseases, 9^th^ Revision, Clinical Modification (ICD-9-CM) code indicates they experienced an injury to their torso, and zero otherwise, for years 2009-2014. For years 2015 and 2016, we instead used the ICD-10 code. Torso injury was derived based on Barell Matrix body regions [39] and was defined as hospital diagnosed injury to the chest (thorax), abdomen, pelvis & urogenital region, trunk, back and buttock, or hip.

Subjects could belong to multiple data sets, however each outcome was analyzed separately, therefore we did not need to account for overlap between data sets in the analysis.

Our primary explanatory variables were seat belt use and occupant age. Seat belt use was coded as a binary variable. Age was categorized into four groups. As our focus is primarily on older occupants, we used more granular categories for older ages, specifically 16-30, 31-64, 65-74, and 75 years or older.

### Statistical Analysis

We estimated the relative risks of the outcomes using conditional Poisson regression, or equivalently, Cox proportional hazards regression with constant survival time and stratified by vehicle [34]. The model was fit via the PHREG procedure in SAS Ver. 9.4 [40]. Crucially, the data likelihood for the models relied only on information from vehicles in which one or more occupants experienced the outcome, and not from the unobserved pairs in which no occupant experienced the outcome. This feature is leveraged to yield unbiased estimates of relative risk associated with seat belt use. By matching the subjects be vehicle, estimates of seat belt protection are conditioned on the subjects having the same crash-and vehicle level factors, such as crash severity and direction, weather conditions, and emergency vehicle response time. Additional details on estimating relative risk using matched cohort data is available in works by Cummings et al. [34, 41].

Age-based relative risks of the outcomes by seat belt use were estimated through an interaction term between age and restraint use. We reported both adjusted and unadjusted relative risks, with adjusted relative risks controlling occupant-level factors that could not be accounted for through the study design. Specifically, the model included indicator variables for occupant sex (male vs. female), seating position (driver seat, right-front passenger seat, or rear passenger seat), and airbag deployment (yes vs. no).

Each of the five linked data sets were treated as “imputed” data sets, and their corresponding estimates were combined into a single estimate via the MIANALYZE procedure in SAS. In doing so, the standard errors corresponding to our relative risk estimates propagate additional uncertainty due to probabilistic linkage. To maintain subject privacy, individual data were not shared between sites. Rather, each site analyzed their data individually using a common SAS script. Results were then shared and combined via a fixed effects meta-analytic approach.[42]

## RESULTS

The fatal injury data set consisted of 12,746 individuals on average (5,571 from Maryland, 4,735 from Ohio, 1,442 from Kentucky, 997 from Utah); the MAIS3+ injury data set consisted of 43,664 individuals on average (21,780 from Maryland, 15,275 from Ohio, 4,362 from Kentucky, 2,247 from Utah); and the torso injury data set consisted of 218,626 individuals on average (93,184 from Maryland, 89,683 from Ohio, 27,308 from Kentucky, 8,451 from Utah). Variability in sample sizes were due to (i) state populations; (ii) types of hospitals included in the data set; (iii) cleanliness and comprehensiveness of PAR data, which affected the success of the linkage procedure; and (iv) years of data available.

Table 1 presents the percentage of occupants in the fatal injury, MAIS3+ injury, and torso injury data sets sustaining the respective injuries. Due to the matched pairs study design, approximately 50% of subjects in their respective data set experienced the outcome. For all three injury types, there is an apparent gradient relationship between higher age and higher percentage sustaining injuries.

**Table 1:** Percentages of injuries sustained among subjects in the fatal, MAIS3+, and torso injury data sets by age and restraint use. ^a^Because sample sizes can vary due to probabilistic linkage, *n* refers to the average sample size across the five data sets that were generated.

| Age | Belt Use | Fatal Injury<br>(*n* = 12,746)<sup>a</sup> | MAIS3+ Injury<br>(*n* = 43,664)<sup>a</sup> | Torso Injury<br>(*n* = 218,626)<sup>a</sup> |
| --- | --- | --- | --- | --- |
| All | Any | 51.2 | 49.5 | 48.7 |
| 16-30 | Any | 47.1 | 45.2 | 44.9 |
| 31-64 | Any | 50.7 | 51.0 | 51.1 |
| 65-74 | Any | 55.7 | 55.2 | 54.0 |
| 75+ | Any | 67.7 | 62.4 | 57.0 |
| All | Belted | 45.1 | 45.2 | 49.9 |
| All | Unbelted | 58.1 | 55.7 | 43.5 |
| 16-30 | Belted | 38.1 | 39.2 | 46.1 |
|  | Unbelted | 54.1 | 51.4 | 40.8 |
| 31-64 | Belted | 42.6 | 44.7 | 51.8 |
|  | Unbelted | 58.9 | 59.7 | 47.2 |
| 65-74 | Belted | 49.8 | 52.8 | 54.6 |
|  | Unbelted | 71.3 | 65.6 | 47.6 |
| 75+ | Belted | 63.8 | 60.9 | 57.9 |
|  | Unbelted | 82.4 | 72.8 | 48.6 |

Table 2 presents age-based relative risks (RR) of each injury outcome associated with seat belt use. The adjusted estimates corresponding to fatal injuries indicate that seat belts are strongly associated with lower risk of fatal injuries, and there is little evidence that the relative risks vary by age. In contrast, while seat belt use was associated with lower risk of MAIS3+ injury for all age groups, the association attenuated with age. Among the youngest age group, seat belt use was associated with 66% lower risk of MAIS3+ injury (RR 0.34, 95% CI 0.30, 0.38), whereas among those aged 75 and older, seat belt use was associated with 38% lower risk of MAIS3+ injury (RR 0.62; 95% CI 0.45, 0.86). The attenuations in effectiveness are illustrated in Figure 1, which plots the adjusted relative risks with 95% confidence limits of fatal injury (Figure 1a), MAIS3+ injury (Figure 1b), and torso injury (Figure 1c)

**Table 2:** Unadjusted and adjusted relative risks of fatal, MAIS3+, and torso injury associated with belt use for the full samples and broken up by age groups. Asterisks denote that there is age-based variability in the association between restraint use and the outcome.

| Outcome | Age Group | Unadjusted RR: belted<br>vs. unbelted | Adjusted RR: belted<br>vs. unbelted |
| --- | --- | --- | --- |
| Fatal Injury | All | 0.27 (0.24, 0.31) | 0.26 (0.23, 0.30) |
|  | 16-30 | 0.26 (0.22, 0.32) | 0.25 (0.21, 0.31) |
|  | 31-64 | 0.26 (0.21, 0.31) | 0.26 (0.21, 0.32) |
|  | 65-74 | 0.25 (0.16, 0.38) | 0.27 (0.17, 0.42) |
|  | 75+ | 0.35 (0.23, 0.53) | 0.35 (0.23, 0.55) |
| MAIS3+* | All | 0.36 (0.33, 0.40) | 0.36 (0.33, 0.39) |
|  | 16-30 | 0.33 (0.30, 0.37) | 0.34 (0.30, 0.38) |
|  | 31-64 | 0.28 (0.25, 0.31) | 0.36 (0.31, 0.41) |
|  | 65-74 | 0.42 (0.31, 0.56) | 0.44 (0.33, 0.60) |
|  | 75+ | 0.61 (0.44, 0.83) | 0.62 (0.45, 0.86) |
| Torso Injury | All | 0.90 (0.85, 0.94) | 0.84 (0.80, 0.89) |
|  | 16-30 | 0.85 (0.80, 0.90) | 0.81 (0.76, 0.87) |
|  | 31-64 | 0.89 (0.84, 0.95) | 0.84 (0.78, 0.90) |
|  | 65-74 | 0.97 (0.79, 1.20) | 0.93 (0.75, 1.15) |
|  | 75+ | 1.10 (0.87, 1.39) | 1.07 (0.84, 1.36) |

**Figure 1:**
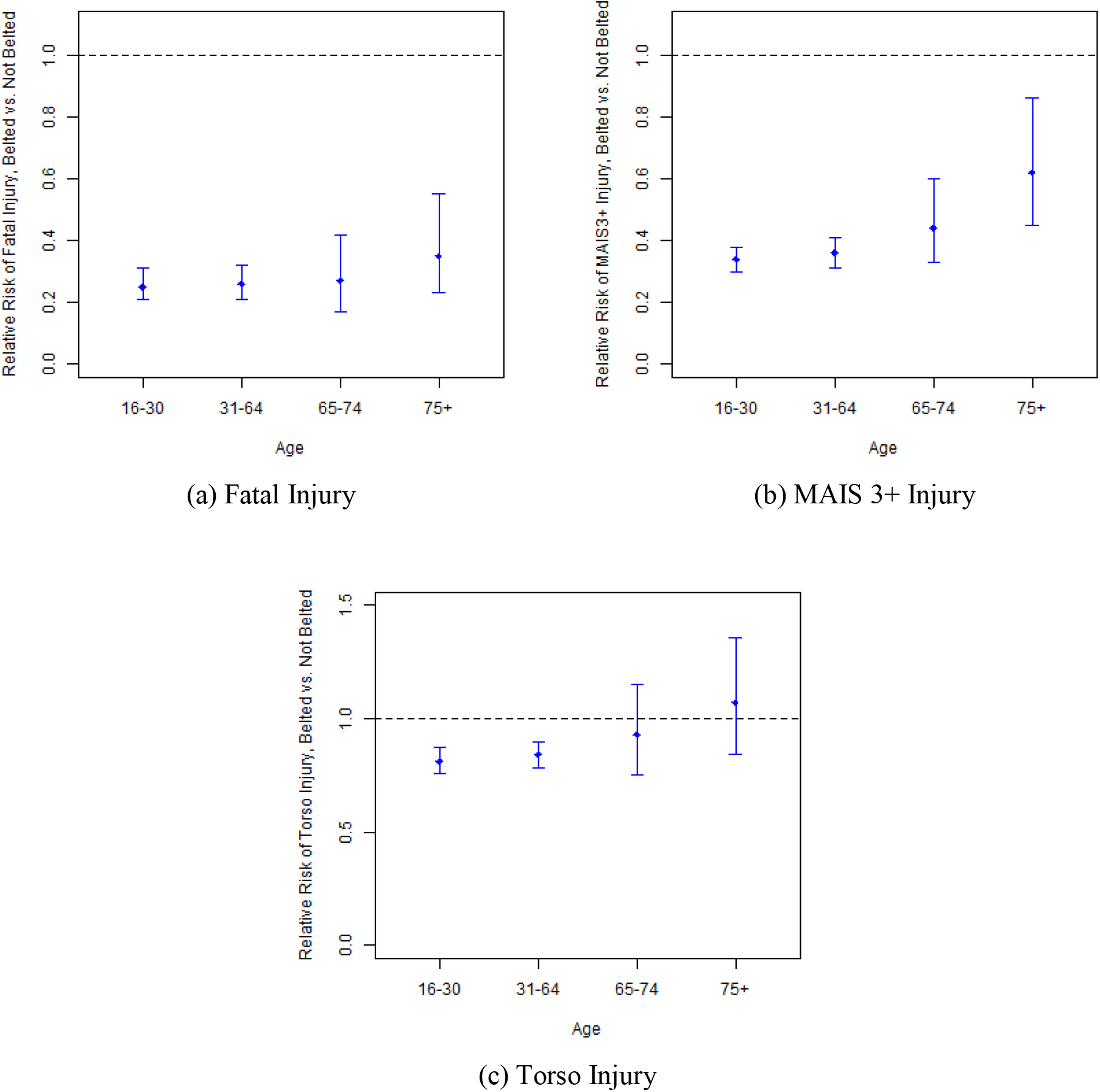
Adjusted relative risks and confidence intervals of fatal injury (a), MAIS3+ injury (b), and torso injury (c) for belted vs. non-belted occupants by age group.

The association between restraint use and torso injury also appeared to attenuate with age (Figure 1c). In the youngest age groups (aged 16-30 and 31-64), seat belt use was associated with lower risk of torso injury (RR = 0.81; 95% CI 0.76, 0.97 for ages 16-30; RR = 0.84; 95% CI 0.78, 0.90 for ages 31-64). However, for the older two age groups, there not evidence to suggest that seat belt use or non-use was associated with risk of torso injury (RR = 0.93; 95% CI 0.75, 1.15 for ages 65-74; RR = 1.07; 95% CI 0.84, 1.36 for ages 75+).

## DISCUSSION

Using data from the CODES database, we estimated age-based associations between seat belt use and fatal, MAIS3+, and torso injuries. To our knowledge it is the first study of US data that examines age-based variability in seatbelt protection against MAIS3+ and torso injuries, as well as one of the first to use leverage hospital-diagnosed injuries for this purpose. As with other studies, we found strong evidence that seat belts are protective for fatal and MAIS3+ injuries. There was also some evidence that restraints were protective for torso injuries among younger occupants. However, seat belt protection against MAIS3+ and torso injuries attenuated with age. Our results did not find evidence that seat belts were protective against torso injuries for those aged 65 and older.

The main strengths of our study were twofold. The first was the quality and scope of the data: our outcomes were primarily derived from hospital records rather than police narrative, and our data were not limited to only fatal crashes. Because seat belt benefit attenuates with crash severity [43], using FARS to estimate seat belt protection against MAIS3+ or torso injuries could have resulted in biased estimates of relative risk. The second strength was the study design. Unlike publicly available data sets such as Crash Injury Research and Engineering Network and the Crash Investigation Sampling System, CODES had enough data that we could leverage a matched cohort design without sacrificing statistical precision. This design was used to control for potential confounding due to all crash- and vehicle-level factors, including those that could not easily by recorded in a crash report. Furthermore, by using matching to control for these factors rather than adjusting for them in a regression model, we did not need to impose a linear relationship between the outcomes and vehicle- and crash-level factors, which may not have accurately characterized their true relationships.

Our study contributes to the already strong evidence in support of seat belts as an effective means of reducing fatalities and injuries in occupants of motor vehicle crashes. We found that seat belts were associated with 74% lower risk of fatality overall. This is consistent with previous studies of restraint use and fatality, many of which were included in a meta-analysis by Høye (2016) [17], whose summary effect indicated that seat belts were associated with 65% lower risk of fatal injury. It is noteworthy that, in contrast to Kahane (2013), we did not find evidence that seat belt protection against fatal injuries attenuated with age [14]. This may be due to our study consisting of newer cars on average, with which have more modern seat belt technology that is effective for older adults [44]. We also found that seat belts were associated with 64% lower risk of MAIS3+ injury, which fell within confidence interval for the summary effect in a meta-analysis by Fouda Mbarga et al. (2018) [16].

The critical finding of this work was that seat belt protection against MAIS3+ injury and torso injuries attenuated with age. Our findings with respect to MAIS3+ injuries contrasted with Noh et al., whose estimated odds ratio associated with failure to wear a seat belt was highest among those aged 65 and older. We found the opposite, in that seat belts exhibited the strongest protection against MAIS3+ injuries among younger occupants. Echoing previous calls to action by, for example, Carter et al. (2014) [3], the implications of our findings with respect to MAIS3+ and torso injury are twofold. First, our results, like studies that found age-based variability in injury severity by age, underscore the need for accurate human conceptual models for all ages and body types. This assertion has been supported by studies of belt positioning [45, 46], body scans [47], and comparison of post-mortem subjects to crash-test dummies [48]. Second, injury prevention systems in motor vehicles should continue to be refined towards vulnerable populations such as the elderly.

It is likely that the primary reason for the age-based attenuation in seat belt protection against MAIS3+ and torso injuries is that elderly occupants are frailer than their younger counterparts [15]. As people age, both bone morphology and the mineral composition of their bones change, with older bones being stiffer but more brittle [49]. These changes mean that older individuals have lower tolerance to deflect forces from the seat belt that lead to torso injuries [50]. Non-epidemiological studies offer additional insight into why age-based variability in restraint effectiveness may exist. Bohman et al. (2019) found that posture changes due to aging can lead to suboptimal belt [45]. Similarly, Brown et al. (2017) found that discomfort due to musculoskeletal morbidities in elderly motor vehicle occupants led to repositioning and use of external seat accessories [51]. Because repositioning can lead to suboptimal belt fit, Brown and colleagues underscored the importance of elderly adults being aware of proper belt fit. Innovations in seat belt design [52, 53] and greater penetration of pretensioners and load limiters [44, 54-56], may also improve torso and other injury outcomes in older adults.

### Limitations

Our study had several limitations. First, torso injury could only be recorded for subjects whose PAR was linked to a hospital record. It is possible that, through unsuccessful linkage or failure to seek medical attention, some subjects with torso injuries were not coded as such. However, because we identified torso injuries using medical evaluation at the hospital, our definition of torso injury is likely more accurate/complete than studies utilizing only data derived from police narrative. Second, our data were drawn from only four states, and were not representative of the entire United States. While we have little reason to believe that true seat belt effectiveness varies between states, additional data from other states would have improved our study. However, on balance, to our knowledge no set of linked crash and hospital data have national scope (except for FARS, which only includes fatal crashes), and a strength of our data is that they draw from states representing three out of four United States Census regions, providing a variety of geographic of geographic and urban/rural driving environments. Third, seat belt use was only available as a binary variable. We could not assess proper use, nor if the vehicle was equipped with technology such as load limiters and pretensioners, which were installed in all US vehicles made in 2008 or later [44].

## CONCLUSIONS

Our study suggests that elderly occupants do not receive the same seat belt benefits as their younger counterparts for MAIS3+ and torso injuries. Although the observed attenuation in injury protection may be largely attributed to elderly occupants’ greater frailty, we encourage that injury prevention measures be tailored to vulnerable populations such as the elderly. Injury outcomes in older occupants may be improved through a combination of greater awareness of proper positioning, continued penetration of modern belt technology, and research on innovative restraint designs.

## Data Availability

The data that support the findings of this study were provided from various entities. They contain protected health information and cannot be made publicly available. Ohio data were provided by the Ohio Department of Public Safety and Ohio Hospital Association, but restrictions apply to the availability of these data, which were used under license for the current study, and so are not publicly available.

## Abbreviations

MVC: Motor Vehicle Crash
CODES: Crash Outcome Data Evaluation System
PAR: Police Accident Report
FARS: Fatal Analysis Reporting System
ICD: International Classification of Diseases
MAIS: Maximum Abbreviated Injury Scale
RR: Relative Risk
CI: Confidence Interval

## Acknowledgements

The authors sincerely thank Ann Nwosu, MS, for her efforts in developing the CODES data set and for her contributions to this project. We would like to acknowledge the contribution of Timothy Kerns, PhD, Cynthia Burch, MS and Shiu Ho, MS to the CODES project in MD.

